# ACQuA: Arrhythmia Classification with Quasi-Attractors

**DOI:** 10.1101/2022.08.31.22279436

**Authors:** William Rudman, Jack Merullo, Laura Mercurio, Carsten Eickhoff

## Abstract

In recent years, deep learning has redefined algorithms for detecting cardiac abnormalities. However, many state of the art algorithms still rely on calculating handcrafted features from a given heart signal that are then fed into shallow 1D convolutional networks or transformer architectures. We propose ACQuA (Anomaly Classification with Quasi Attractors), a task agnostic algorithm that can be used in a wide variety of cardiac settings, from classifying cardiac arrhythmias from ECG signals to detecting heart murmurs from PCG signals. Using theorems from dynamical analysis and topological data analysis, we create informative attractor images that 1) are human distinguishable and 2) can be used to train small, off the shelf deep neural networks for anomaly classification. In the George B. Moody 2022 Challenge, we receive an official score of 0.433 (263/305) for murmur classification and a score of 12616 (208/305) for outcome classification. Additionally, we evaluate our model on the CinC 2017 Challenge data that tasks practitioners to classify cardiac arrhythmias from ECG signals. On the CinC 2017 Challenge data, we improve upon the winning F1 scores by approximately 14% on the hidden validation data.

## 1. Introduction

Improving methods to detect cardiovascular disease can both improve patient care and reduce the economic burden that untreated cardiac abnormalities place on the healthcare system. Traditional approaches to cardiac arrhythmia classification and murmur classification input handcrafted features of a ECG/PCG signals into a deep learning architecture- typically a transformer or 1D-CNN. We propose a novel approach that identifies cardiac abnormalities using the quasi-attractor of an ECG/PCG signal. The quasi-attractor of a signal is the approximate set of steady states that the underlying dynamical system governing the signal converges to over time. Analysis of quasi-attractors is widely used for time-series classification and forecasting in dynamical analysis. However, current analysis of quasi-attractors requires an expensive post-processing step that involves calculating features of the geometry of the quasi-attractor. Instead, we input images of the quasi-attractor into a ResNet model to automatically extract the most relevant features for classification. As our model is a general framework for signal classification, we test our model on two common tasks: cardiac arrhythmia classification from ECG signals and murmur detection from PCG signals. Although ACQuA performs poorly in detecting murmurs from PCG signals in the George B. Moody Physionet 2022 Challenge, our model outperforms several top baselines for cardiac arrhythmia classification from the CinC/Physionet 2017 Challenge data. The remaining sections of our paper will discuss 1) the data pre-processing and pre-training steps, 2) detail our quasi-attractor imaging methods, 3) specify model hyperparemeters, and; 4) present results for murmur detection and cardiac arrhythmia classification. We conclude our paper with a thorough analysis of the success and failure cases of ACQuA.

## 2. Methods

### 2.1. Pre-Processing & Pre-Training

Given the interpatient variability of cardiac rhythms and the inherently noisy nature of cardiac signals, several preprocessing steps are required. Firstly, we apply a low-pass butterworth filter to remove low frequency noise from the signal. Next, we re-sample the signal to a frequency of 1000Hz and lastly, we normalize bounds such that the signals are contained in the interval [0, 1].

The CinC 2017 challenge data contains 8,528 ECG signals recorded on an AliveCor device which generates a single channel ECG signals for a patient. There are four classes present in the CinC 2017 data: Normal, AFib, Other and too noisy to be classified. Given the relatively large size of the dataset and the fact we can use a single-channel ResNet architecture, we can train ACQUA with no additional pre-training steps.

For the George B. Moody PhysioNet 2022 Challenge, we pre-trained ACQUA on the CinC 2016 Challenge data. The CinC/PhysioNet 2016 data contains 3,126 heart sound recordings from a single auscultation location and tasks challengers to determine whether or not a murmur is present in the recording. In order to pre-train ACQuA on the CinC 2016 data, we have to input a stack of 5 duplicates of the model since the patients in the George B. Moody PhysioNet 2022 Challenge data can have up to five PCG recordings obtained from different locations.

### 2.2. Attractor Imaging

Time-Delay Embeddings are the most commonly used method for quasi-attractor reconstruction. Let {*x*_0_, *x*_1_, …, *x*_*N*_*}* be a collection of points sampled from a real-valued function *f* at equal intervals. We define the *time-delay embedding* of *f* centered at *x*_0_, to be the following set of points contained in ℝ ^*M*^ :

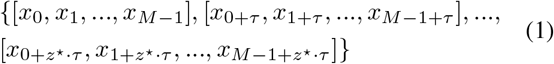

where *M* denotes the *embedding dimension, τ* denotes the *time delay* and 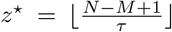. We use ⌊·⌊ to denote the floor function. The generalized Taken’s Embedding Theorem [1] states that there exists a choice of *M* and *τ* such that the time-delay embedding is sampled from a smooth manifold diffeomorphic (i.e. there exists an isomorphism between the two smooth manifolds) to the true attractor of the underlying system. Therefore, the quality of the time-delay embedding is dependent on the choice of hyperparameters *M* and *τ*.

Currently, there is no definitive way of choosing optimal parameters *M* and *τ* when creating time-delay embeddings. The most common practices for choosing *M* and *τ* are determined by either autocorrelation or mutual information methods to choose *τ* and false nearest neighbor algorithms to choose *M* [2]. However, many authors naïvely select *M* = 3 and *τ* = 1 which may not capture the true dynamics of the underlying system. The time-delay embedding more accurately models the true attractor of the dynamical system when the embedding dimension is chosen to be larger than or equal to the length of the period of the underlying system [3].

For the CinC 2017/CinC 2022 Challenge data, we set *τ* = 1 and *M* = 45, 60, respectively. These parameters have been determined by performing small-scale experiments by altering *M* and *τ*. Using an embedding dimension greater than 3 requires further techniques to make attractor imaging tractable. Kim et al. [4] prove that performing PCA on time-delay embeddings reduces topological noise that can otherwise alter the geometry of the quasi-attractor. After obtaining the time-delay embedding of an ECG signal, we perform PCA to reduce the dimension of the quasi-attractor to 2. For the CinC 2022 data, we create attractor images for every lead present in the data and stack the image tensors before inputting them into a ResNet-18 model where each lead corresponds to an input channel.

### 2.3. Model Details

We use a custom ResNet model to perform image classification on the quasi-attractor images of both PCG and ECG signals. The ResNet family of models is a commonly used architecture in image classification and has achieved state-of-the-art results in several computer vision tasks. The residual skip connections that give ResNet its namesake offer a significant advantage over traditional 2D CNNs as ResNet allows for the training of deeper networks and mitigates the impact of vanishing gradients. We select a ResNet architecture to keep our model computationally efficient and mobile. We train our model using binary cross entropy loss for 50 epochs with a learning rate of 0.001 and a batch size of 40. Note that we create an attractor for each of the 5 possible auscultation location. Before inputting the images into ResNet, we stack the signal tensors so that each signal corresponds to an input channel in the network. We modify Resnet-18 to accept more than 3 (typically the red, blue, and green pixels) channels. If a patient is missing a recording from a given location, we pad the tensors with an image of all 0s. Both the murmur classifier and the outcome classifier share weights of the ResNet image encoder and the overall model is trained with an evenly-weighted joint training objective. Figure 3 illustrates the ACQuA model architecture used in the George B. Moody 2022 challenge. For the CinC/Physionet 2017 Challenge data, the AliveCor device produces a single channel image, grayscale image– simplifying the overall model architecture. Namely, for the CinC/Physionet 2017 data, we use a one channel version of the ResNet model used in our ACQuA model pipeline.

**Figure 1.**
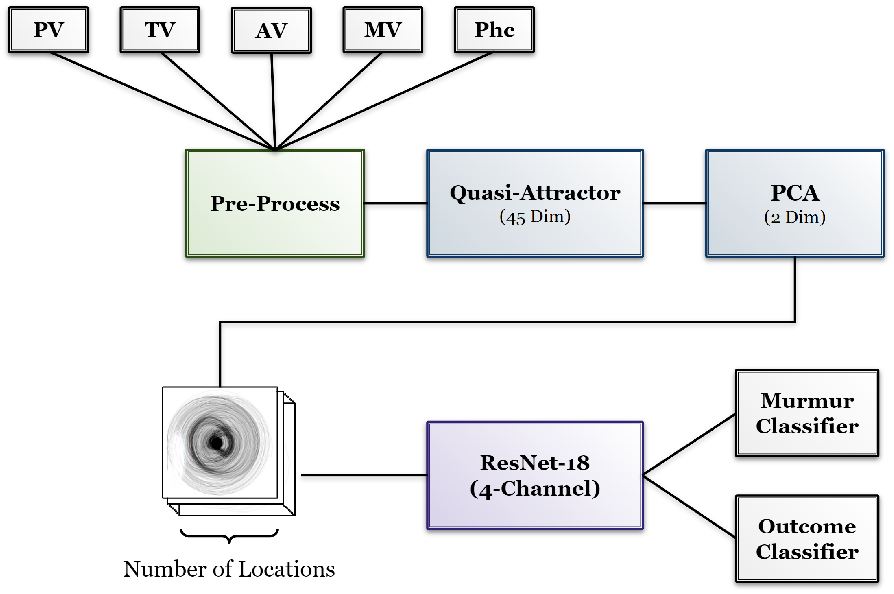
Illustration of the ACQuA model pipeline. Cardiac signals recorded at different locations are converted to images through PCA on an attractor imaging analysis and fed into a custom ResNet architecture.

**Figure 2.**
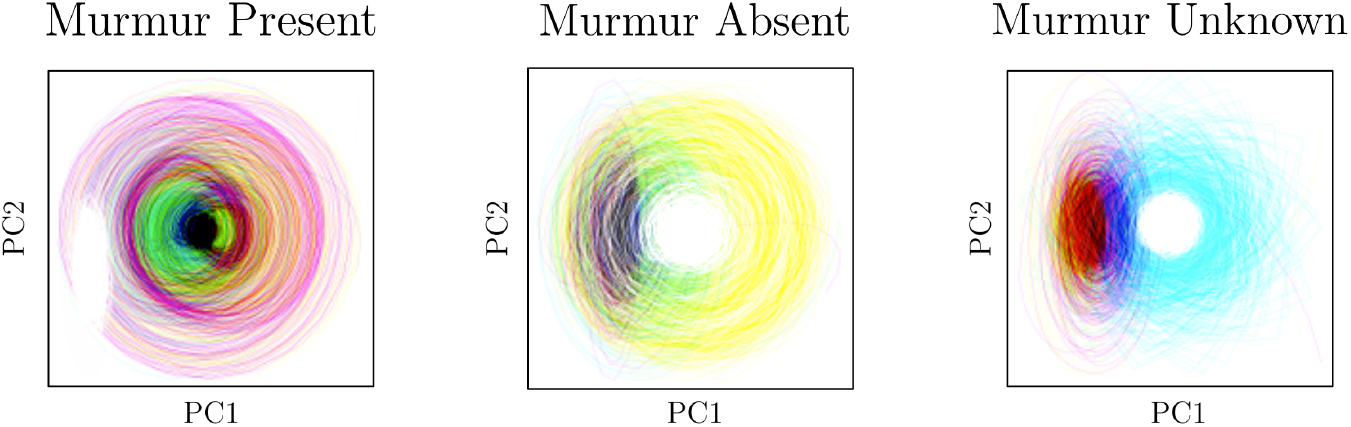
Sample quasi-attractor images for present, absent and unknown murmur classes from George B. Moody 2022 Challenge data. Note, that we represent stack the channels to create a single RBGA image from the different auscultation locations.

**Figure 3.**
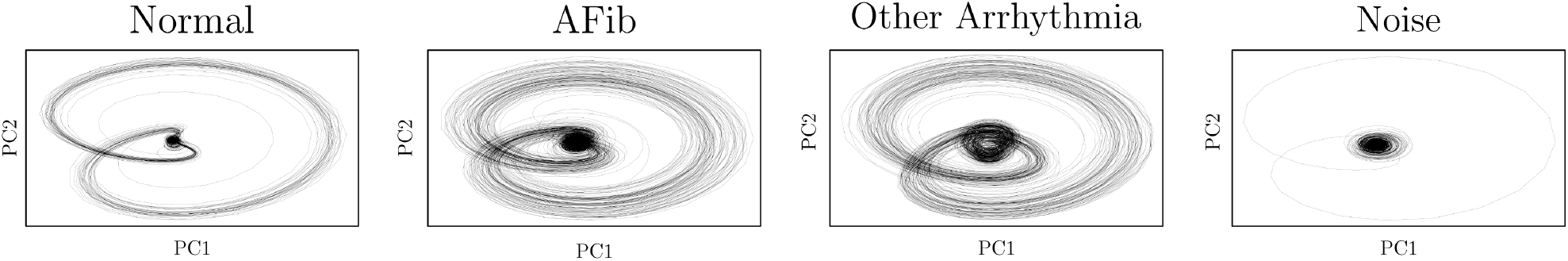
Sample quasi-attractor images for ECG signals for the Normal, AFib, Other and Noise classes from the CinC/Physionet 2017 challenge data.

## 3. Results

In this section, we report both ACQuA’s performance on the George B. Moody Challenge validation data and ACQUA’s performance on the CinC/Physionet 2017 Challenge. Given that the CinC 2017 Challenge test data is not publicly available, we report average F1 scores on the validation data and compare results with the top three papers from the challenge. Figure 4 details our results on the George B. Moody 2022 Challenge.

**Figure 4.**
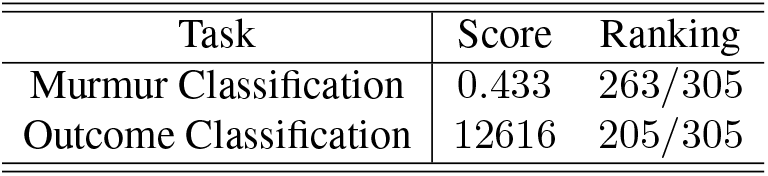
Results of ACQuA on the George B. Moody 2022 Challenge.

**Figure 5.**
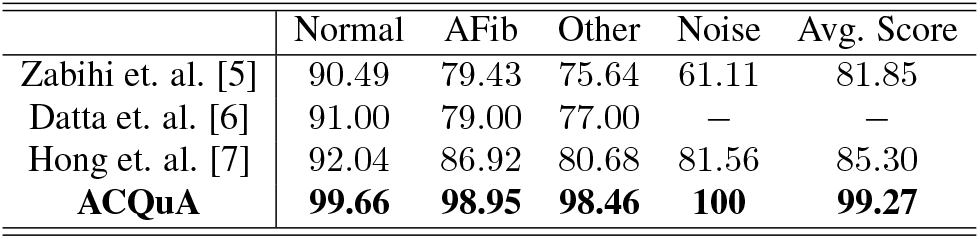
CinC/Physionet 2017 Challenge *F*_1_ scores on the validation data.

Table 5 demonstrates the efficacy of ACQuA in detecting and diagnosing cardiac arrhythmias from a single channel. Our method improves upon the CinC 2017 Challenge winner by approximately 14% with our model scoring a perfect 100% F1 on the Noise class where many of the top performing algorithms struggle to exceed an F1 score of 81.56%.

## 4. Discussion

In this paper we present our novel pipeline ACQuA: Anomaly Classification with Quasi Attractors. ACQuA is a flexible framework that can be applied to a variety of classification tasks from diagnosing cardiac arrhythmias from ECG signals and detecting heart murmurs from PCG signals. In addition to competing in the George B. Moody 2022 Challenge, we evaluate our methods on the CinC/Physionet 2017 challenge.

The performance of our algorithm differs drastically depending on the task. On the CinC/Physionet 2017 challenge data, ACQuA improves upon the winning algorithms, however, our model performs poorly on identifying murmurs from PCG signals in the George B. Moody 2022 Challenge. We hypothesize that the discrepancy in the performance between the two tasks is due to the following factors: 1) the number of training/pre-training samples, 2) a lack of consistency in the number of auscultation location per patient, and; 3) high inner-class variability in quasi-attractors obtained from PCG signals.

The backbone of ACQuA is a ResNet model with approximately 16-million parameters. Given the number of parameters in the ResNet model, training ACQuA to detect heart murmurs from heart sounds obtained from 942 patients is extremely challenging. Pre-training with the CinC/Physionet 2016 Challenge data (3,126 recordings) did lead to an increase in performance, however, we had to make several adjustments in order to re-format the challenge data to be input into our ACQuA architecture. The CinC/Physionet 2016 Challenge data contains heart sounds from a single location where the George B. Moody 2022 Challenge data contains up to five different locations for a single patient. In order to pre-train our model, we had to input 5 duplicate signals into each of the 5 channels of the ResNet Model. Meaning that in the course of pretraining each channel received information from a variety of different auscultation locations, however, in training on the 2022 data each input channel received information from a single auscultation location. Namely, during training channel 1 only receives heart sounds obtained from the pulmonary vale, channel 2 only receives recordings from the aortic valve and so on. We experimented with concatenating signals from every auscultation location obtained for a patient into a single signal and then computing the quasi-attractor. Unfortunately we were unable to evaluate our method on the hidden validation data due to errors in our code.

Quasi-attractor images created from PCG signals vary wildly within class labels. Given the complexity of murmur representations in quasi-attractor images, a wealth of data would be needed for our model to ascertain patterns between quasi-attractor images where a murmur is present and images where a murmur is absent. The primary reason for the high inner-class variability is that the number and location of signals varies between each patient. A large part of the success of ACQuA in cardiac arrhythmia classification is due to the uniqueness of the quasi-attractor images between different arrhythmia types and the interpatient consistency within each class.

## 5. Conclusion

In this paper we present a novel architecture, ACQuA (Anomaly Classification with Quasi-Attractors) for classifying cardiac abnormalities. Our approach leverages advances in image classification for the purpose of effectively classifying cardiac time-series signals. We use the CinC/Physionet 2017 data to classify arrhythmias from an AliveCor device and compete in the George B. Moody 2022 challenge to evaluate the efficacy of our model in detecting the presence of heart murmurs from PCG recordings. Results show one successful use case (ECG classification) and one unsuccessful use case (Murmur identification) of our pipeline. We propose a several reasons for the performance differential in Section 4, however, more work is needed to investigate the reasons behind the discrepancy in performance between ECG and PCG classification tasks. Although our model performed poorly on the murmur identification and outcome classification tasks present in the George B. Moody 2022 challenge, ACQuA excels in cardiac arrhythmia classification. More work is needed to determine which features of a task allow our approach to succeed; we are excited by future research in this direction as well as identifying other tasks that can benefit from quasi-attractor image classification.

## Data Availability

The George B. Moody PhysioNet 2022 Challenge data is available at: https://moody-challenge.physionet.org/2022/
The CinC Physionet 2017 Challenge data is available at: https://archive.physionet.org/physiobank/database/challenge/2017/

https://archive.physionet.org/physiobank/database/challenge/2017/

## Acknowledgements

This research is supported in part by the NSF (IIS-1956221)T32 GM128596). The views and conclusions contained herein are those of the authors and should not be interpreted as necessarily representing the official policies, either expressed or implied, of NSF, NIH, or the U.S. Government.

## References

[1] Takens F. Detecting Strange Attractors in Turbulence. Lecture Notes in Mathematics, volume 898. ISBN 978-3-540-11171-9, 11 2006; 366–381.

[2] Krakovska A, Mezeiova K, Budacova H. Use of false nearest neighbours for selecting variables and embedding parameters for state space reconstruction. Journal of Complex Systems 03 2015;2015:12.

[3] Perea J, Harer J. Sliding windows and persistence: An application of topological methods to signal analysis, 2013.

[4] Kim K, Kim J, Rinaldo A. Time series featurization via topological data analysis, 2019.

[5] Zabihi M, Rad AB, Katsaggelos AK, Kiranyaz S, Narkilahti S, Gabbouj M. Detection of atrial fibrillation in ecg handheld devices using a random forest classifier. In 2017 Computing in Cardiology (CinC). 2017; 1–4.

[6] Datta S, Puri C, Mukherjee A, Banerjee R, Choudhury AD, Singh R, Ukil A, Bandyopadhyay S, Pal A, Khandelwal S. Identifying normal, af and other abnormal ecg rhythms using a cascaded binary classifier. In 2017 Computing in Cardiology (CinC). 2017; 1–4.

[7] Hong S, Wu M, Zhou Y, Wang Q, Shang J, Li H, Xie J. Encase: An ensemble classifier for ecg classification using expert features and deep neural networks. In 2017 Computing in Cardiology (CinC). 2017; 1–4.

